# BNT162b2 induced neutralizing and non-neutralizing antibody functions against SARSCoV-2 diminish with age

**DOI:** 10.1101/2022.08.12.22278726

**Authors:** Timothy A. Bates, Pei Lu, Ye jin Kang, Devin Schoen, Micah Thornton, Savannah K. McBride, Chanhee Park, Daehwan Kim, William B. Messer, Marcel E. Curlin, Fikadu G. Tafesse, Lenette L. Lu

**Author notes:** Correspondence: Lenette Lu MD PhD, UT Southwestern Medical Center, 5323 Harry Hines Blvd, Dallas, TX 75390, Fikadu G. Tafesse PhD –, Marcel E Curlin MD –. These authors contributed equally (Timothy A. Bates, Pei Lu).

## Abstract

Each novel SARS-CoV-2 variant renews concerns about decreased vaccine efficacy caused by evasion of vaccine induced neutralizing antibodies. However, accumulating epidemiological data show that while vaccine prevention of infection varies, protection from severe disease and death remains high. Thus, immune responses beyond neutralization could contribute to vaccine efficacy. Polyclonal antibodies function through their Fab domains that neutralize virus directly, and Fc domains that induce non-neutralizing host responses via engagement of Fc receptors on immune cells. To understand how vaccine induced neutralizing and non-neutralizing activities synergize to promote protection, we leverage sera from 51 SARS-CoV-2 uninfected health-care workers after two doses of the BNT162b2 mRNA vaccine. We show that BNT162b2 elicits antibodies that neutralize clinical isolates of wildtype and five variants of SARS-CoV-2, including Omicron BA.2, and, critically, induce Fc effector functions. FcγRIIIa/CD16 activity is linked to neutralizing activity and associated with post-translational afucosylation and sialylation of vaccine specific antibodies. Further, neutralizing and non-neutralizing functions diminish with age, with limited polyfunctional breadth, magnitude and coordination observed in those ≥65 years old compared to <65. Thus, studying Fc functions in addition to Fab mediated neutralization provides greater insight into vaccine efficacy for vulnerable populations such as the elderly against SARS-CoV-2 and novel variants.

## Introduction

Neutralizing antibody responses are among the core measures of vaccine efficacy in the COVID-19 pandemic (Garcia-Beltran et al., 2022; Liu et al., 2021). Yet even when neutralization is compromised in the setting of new SARS-CoV-2 variants (Planas et al., 2022) and cases of vaccine breakthrough infections rise, protection from hospitalization remains relatively high (Altarawneh et al., 2022; Collie et al., 2022; Nasreen et al., 2022; Tang et al., 2021). Thus, the continued emergence of new variants highlights the need to understand vaccine efficacy through protection from disease in addition to prevention of infection.

Though one of the key components of immune protection, the complexity of polyclonal antibody responses and its roles in disease remain only partially understood. For SARS-CoV-2, attention has focused on leveraging direct neutralization of virus by antigen recognition via the Fab domain. However, the overall magnitude of neutralizing responses in patients with severe COVID-19 is higher compared to mild disease, suggesting that neutralizing activity alone poorly captures the capacity to protect from serious illness (Lucas et al., 2021; Savage et al., 2021). Independently, data from multiple large clinical trials have demonstrated that convalescent plasma carrying neutralizing activity does not prevent infection or disease in humans (Begin et al., 2021; Group, 2021; Writing Committee for the et al., 2021), suggesting that passive transfer of neutralizing polyclonal antibodies is insufficient to confer protection. These lines of evidence show that in SARS-CoV-2 infection, more nuanced evaluations of neutralizing responses with respect to potency (Garcia-Beltran et al., 2021) and dynamics (Lucas et al., 2021), and immune responses beyond neutralization are vital in understanding pathogenesis.

Antibodies function through the combination of the Fab domain that directs neutralizing activity against microbial targets and the Fc domain that induces non-neutralizing functions (Lu et al., 2018). Through binding Fc receptors expressed on innate and adaptive immune cells as well as activation of complement, antibody Fc domains have the ability to induce a spectrum of host responses directed against an antigen recognized by the Fab domain (Pincetic et al., 2014). Thus, antibody Fc effector functions have the potential to impact outcomes of SARS-CoV-2 infection and protection in vaccines.

Studies using monoclonal antibodies targeting SARS-CoV-2 show that Fc effector functions can be protective. Passive transfer of monoclonal antibodies with mutations that abrogate Fc domain binding to Fc receptors result in increased SARS-CoV-2 viral load and decreased survival in multiple animal models when compared to intact antibodies (Schafer et al., 2021; Suryadevara et al., 2021; Ullah et al., 2021; Yamin et al., 2021). This effect is more pronounced with therapeutic than prophylactic administration (Winkler et al., 2021). Thus, monoclonal antibody Fc functions support neutralizing activity to prevent viral entry. Moreover, even after viral infection, Fc functions can inhibit disease progression.

Conversely, several lines of evidence show that Fc effector functions in polyclonal responses during SARS-CoV-2 infection could be pathogenic. Post-translational IgG glycosylation is altered with disease severity in many ways (Farkash et al., 2021; Petrovic et al., 2021; Vicente et al., 2022) but one consistent observation across several studies is that decreased IgG fucosylation correlates with worsening clinical symptoms and hospitalization (Chakraborty et al., 2021; Chakraborty et al., 2022; Larsen et al., 2021). The proposed mechanism of pathology is through increased binding to the activating Fc receptor FcγRIIIa/CD16a. In an *in vitro* poly I:C stimulated human macrophage model with FcγRIIIa/CD16a expression, addition of afucosylated compared to fucosylated IgG from patients infected with SARS-CoV-2 enhances secretion of the pro-inflammatory cytokine IL-6 (Hoepel et al., 2021; Larsen et al., 2021). In monocytes, FcγR mediated activation can cause pyroptosis (Junqueira et al., 2022). In a human Fc receptor transgenic mouse model, passive transfer of afucosylated polyclonal IgG from individuals with severe COVID-19 increases production of IL-6 and TNFα but not the anti-inflammatory IL-10 (Chakraborty et al., 2022). Consistent with these data, FcγRIIIa/CD16a natural killer (NK) cell activation that leads to antibody dependent cellular cytotoxicity (ADCC) is enhanced with symptom severity and normalizes upon convalescence (Chakraborty et al., 2021). The low affinity activating FcγRIIa/CD32a and inhibitory FcγRIIb/CD32b along with the high affinity FcγRI/CD64 mediate the non-neutralizing Fc effector functions of antibody dependent cellular phagocytosis (ADCP) by monocytes. Neutrophils express antibody receptors for both IgG, the activating high affinity FcγRI, low affinity FcγRIIa and FcγRIIIB, as well as IgA, the low affinity FcαRI. These, along with complement receptors CR1 and CR3 contribute to neutrophil phagocytosis. Finally, C1q binding to IgG and IgM Fc domains activate complement pathways through C3 deposition (Lofano et al., 2018; Peschke et al., 2017; Quast et al., 2015; van Osch et al., 2021). In contrast to FcγRIIIa/CD16a activities, the implications of FcγRIIa/CD32a and FcγRIIb/CD32b mediated phagocytosis and complement activation in SARS-CoV-2 are less clear given the variability in cohort populations with respect to clinical outcomes, demographics and co-morbidities (Adeniji et al., 2021; Bartsch et al., 2021; Herman et al., 2021; Klingler et al., 2021; Selva et al., 2021). However, that multiple Fc effector functions in infection and disease are detectable suggest that these responses if induced by vaccines could influence outcomes.

For COVID-19 vaccines, neutralizing titers are often used to extrapolate protective efficacy (Lustig et al., 2021). While antibody dependent NK cell activation (ADNKA), ADCC, ADCP by monocytes, ADNP by neutrophils and complement activation are also elicited (Alter et al., 2021; Gorman et al., 2021; Kaplonek et al., 2022), it is unclear whether these Fc effector functions are protective, inert, or pathogenic. Moreover, how non-neutralizing antibody functions impact direct neutralization of live virus is not known. To assess the relationships between Fab and Fc domain functions in polyclonal responses from vaccination, we evaluated immune sera from SARS-CoV-2 uninfected health-care workers who received two doses of the BNT162b2 mRNA vaccine. We assessed neutralization against SARS-CoV-2 wildtype virus (WA.1) and five clinical variants: Alpha (B.1.1.7), Beta (B.1.351), Gamma (P.1), Delta (B.1.617.2) and Omicron (BA.2). We measured vaccine specific antibody Fc features of isotype, Fc receptor binding, Fc effector functions and IgG glycosylation. We found heterogeneous neutralizing and non-neutralizing antibody responses. Neutralization across variants correlated with FcγRIIIa/CD16a effector functions in an age but not sex dependent manner. Post-translational afucosylation and sialylation of vaccine specific antibodies associated with enhanced FcγRIIIa/CD16a activity. Neutralizing and non-neutralizing functions independently and collectively diminished with age, limiting polyfunctional breadth, magnitude, and coordination in those ≥65 years old compared to <65. Our results show that assessment of vaccine efficacy against SARS-CoV-2 and novel variants is enhanced by the addition of diverse Fc functions to traditional Fab functions, particularly in vulnerable populations such as the elderly.

## Results

### Study subjects

To evaluate polyclonal antibody responses to mRNA COVID-19 vaccines, sera were collected from 51 adults who received two doses of BNT162b2 vaccine between December 2020 and February 2021 (Table) (Bates et al., 2021a). These individuals spanned a spectrum of ages from 21-82 years. To limit confounding variables, samples were selected to minimize variations in time between vaccine dose 1 and 2 (20-22 days, variation of 2 days) and dose 2 to sample collection (14-15 days, variation of 1 day); sex distribution was balanced. To avoid the complicating factor of hybrid immunity due to SARS-CoV-2 infection, we excluded individuals with report of prior infection or active symptoms and performed confirmatory testing to verify the absence of detectable SARS-CoV-2 nucleocapsid specific antibodies (Supplemental Figure 1A).

**Table:** Demographics and vaccination status

| BNT162b2-vaccinated donors |  |
| --- | --- |
| Characteristic | Total (N=51) |
| Median age | 50 yo (21-82) |
| Sex – no. (%) |  |
| Female | 28 (54.9%) |
| Male | 23 (45.1%) |
| Median time between vaccine doses | 21 days (20-22) |
| Median time between second dose and sample collection | 14 days (14-15) |

### Neutralizing antibody titers of wildtype and SARS-CoV-2 variants

Using the SARS-CoV-2 receptor binding domain (RBD) antigen encoded by BNT162b2 (Vogel et al., 2021), we found that 100% of individuals after two doses of the vaccine had detectable antigen specific IgG compared to 51% with IgA (Supplemental Figure 1A). Thus, consistent with other studies, the primary isotype mediating antibody function two weeks after a second BNT162b2 dose was IgG (Brewer et al., 2022; Collier et al., 2021). To assess direct neutralization, we performed focus reduction neutralization tests using live wildtype SARS-CoV-2 (isolate WA1/2020) virus (Supplemental Figure 1B). Consistent with the generation of RBD specific IgG, all individuals had detectable capacity to neutralizing activity. Linear regression showed that neutralization was dependent on RBD specific antibodies, specifically IgG and not IgA (Figure 1A).

**Figure 1:**
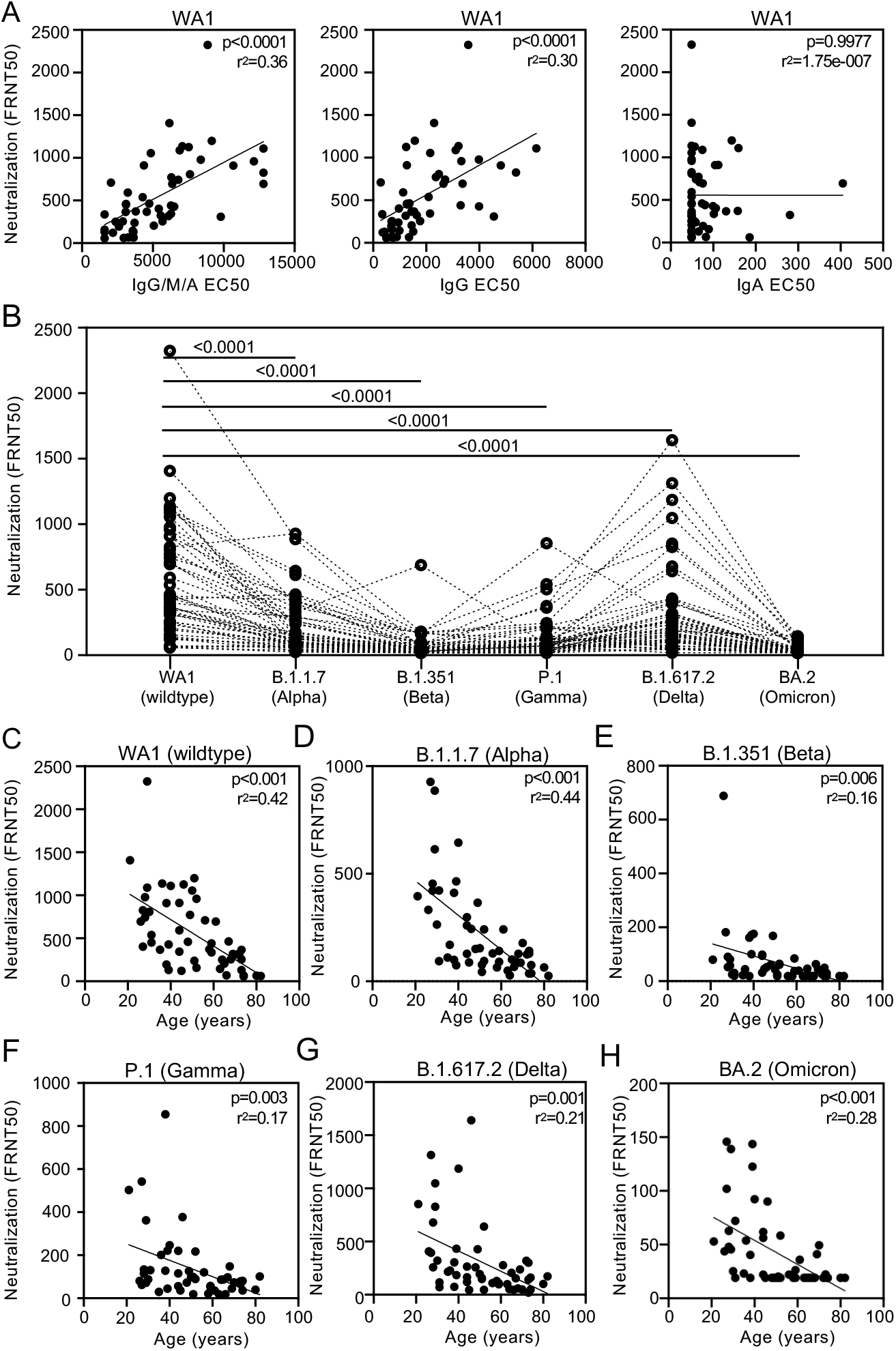
BNT162b2 induced IgG mediates age-dependent neutralization of wildtype and SARS-CoV-2 clinical variants. (A) Live SARS-CoV-2 neutralization (FRNT50) wildtype WA1 and receptor binding domain (RBD) specific IgG/M/A, IgG and IgA EC50 values (Supplemental Figure 1A) are plotted with relationship assessed by linear regression. (B) Neutralization of live SARS-CoV-2 wildtype WA1 and variants (Supplemental Figure 1B) are depicted with each dotted line representing a single individual and statistical significance calculated by Wilcoxon matched pair signed rank test. Live SARS-CoV-2 neutralization (FRNT50) for (C) wildtype (WT) and variants (D) B.1.1.7 (Alpha), (E) B.1.351 (Beta), (F) P.1 (Gamma), (G) B.1.617.2 (Delta) and (H) BA.2 (Omicron) and age in years are plotted with relationship assessed by linear regression and p values adjusted for sex.

We next measured the neutralizing activity of vaccinee sera against SARS-CoV-2 clinical isolates of the viral variants Alpha (B.1.1.7), Beta (B.1.351), Gamma (P.1), Delta (B.1.617.2) and Omicron (BA.2), (Wang et al., 2022) (Supplemental Figure 1B). We used live virus instead of pseudovirus to more effectively model physiological ratios and spectrum of SARs-CoV-2 antigens during infection and replication (Syed et al., 2021). We found that neutralization of variants was diminished relative to wildtype and varied by viral variant and individual (Figure 1B) with the lowest levels detected against Omicron (BA.2), consistent with other studies (Evans et al., 2022; Kurhade et al., 2022; Wang et al., 2022). More specifically, while all individuals had detectable neutralization against wildtype and Alpha (B.1.1.7), only 57% had detectable responses against Omicron (BA.2) which were lower on average than for other variants. While sex can impact immune responses (Scully et al., 2020), we observed no sex based difference in neutralization. (Supplemental Figure 1C). However, we did detect a negative correlation between age and neutralization (Supplemental Figure 1D). To incorporate both age and sex into our evaluations, we used multivariable regression to assess the relationships with neutralization. We found that neutralization of wildtype and variants was negatively correlated with age (Figure 1C-H) but the correlation with sex remained non-significant. Upon review of the 43% of individuals with no detectable neutralizing activity against Omicron (BA.2), we observed that the median age of this subgroup was 63.5 years, above the median age of 50 for all individuals in this study. Consistent with other reports, these data showed that BNT162b2 induced RBD IgG neutralized SARS-CoV-2 wildtype virus and multiple variants in an age but not sex dependent manner (Bates et al., 2021a; Collier et al., 2021; Kawasuji et al., 2021).

### Vaccine specific Fc effector functions

Because IgG was the predominant vaccine specific isotype, we focused on RBD specific IgG effector functions to evaluate the relationship between viral neutralization via by the Fab domain and non-neutralizing Fc activity. We began by measuring RBD specific antibody binding to the activating receptors, FcγRIIIa/CD16a and FcγRIIa/CD32a, and the sole inhibitory receptor, FcγRIIb/CD32b, because engagement of these low affinity Fc receptors are modifiable by dynamic changes in subclass and post-translational glycosylation (Alter et al., 2018; Nimmerjahn and Ravetch, 2005; Pincetic et al., 2014). We found that RBD specific IgG binding to FcγRIIIa/CD16a (Figure 2A), FcγRIIa/CD32a (Figure 2B) and FcγRIIb/CD32b (Figure 2C) positively correlated with SARS-CoV-2 neutralization in varying degrees.

**Figure 2:**
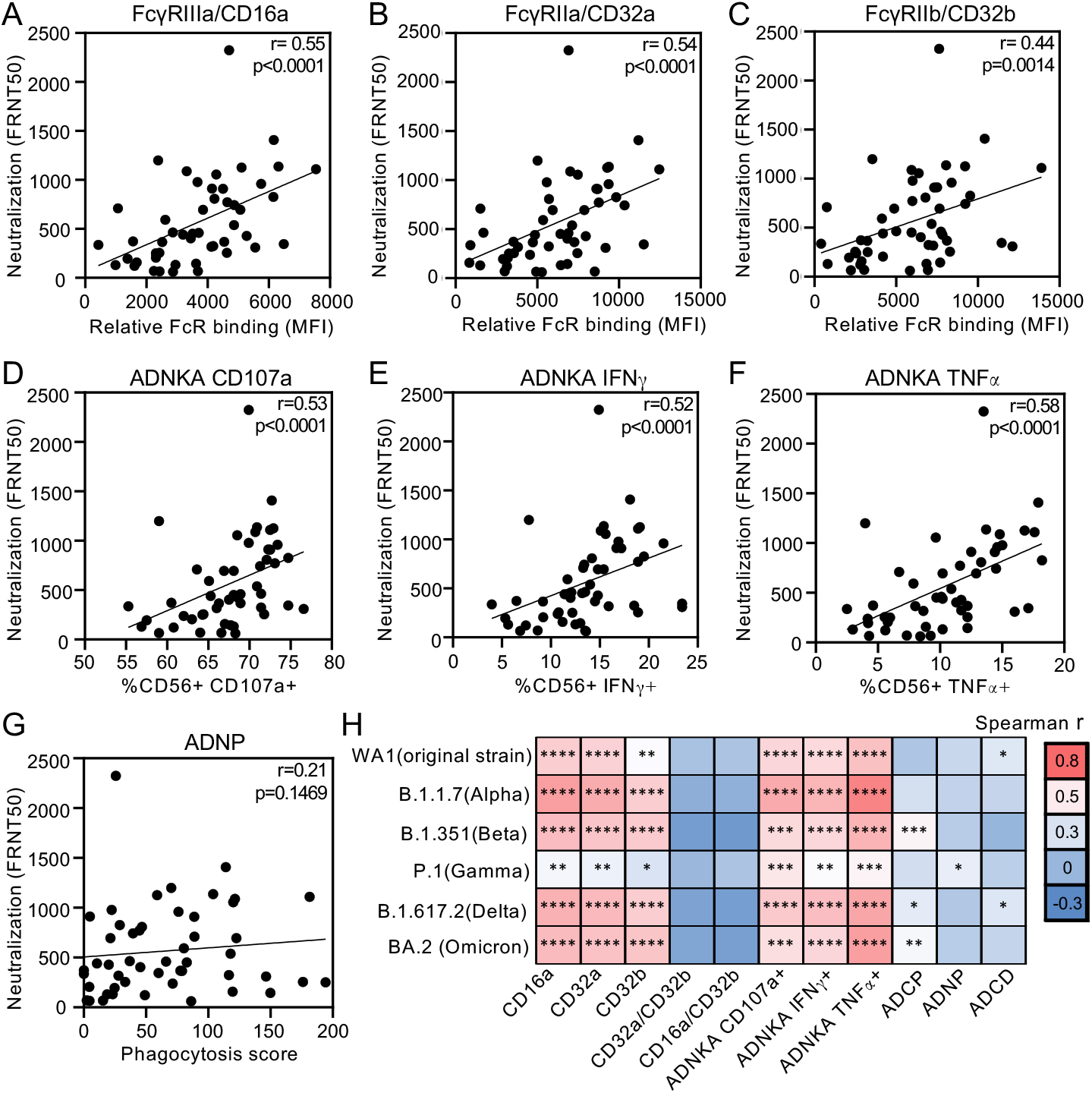
Vaccine specific IgG induction of FcγRIIIa/CD16 effector functions correlate with neutralization of wildtype and SARS-CoV-2 clinical variants. Relationships between live SARS-CoV-2 WA1 neutralization (FRNT50) and receptor binding domain (RBD) specific relative binding to (A) FcγRIIIa/CD16a, (B) FcγRIIa/CD32a and (C) FcγRIIb/CD32b, RBD specific antibody dependent natural killer cell activation (ADNKA) determined by (D) CD107a expression, (E) IFNγ production and (F) TNFα secretion, and (G) RBD specific antibody dependent neutrophil phagocytosis (ADNP) are shown. (H) Heatmap summarizes Spearman correlations (Supplemental Figure 2) between neutralization of SARS-CoV-2 wildtype WA1 and variants with relative binding of RBD specific IgG to activating (FcγRIIIa/CD16a and FcγRIIa/CD32a), inhibitory (FcγRIIb/CD32b) and ratios of activating:inhibitory FcγR (FcγRIIIa/CD16a:FcγRIIb/CD32b and FcγRIIa/CD32a:FcγRIIb/CD32b) binding and Fc effector functions ADNKA, antibody dependent cellular phagocytosis (ADCP), ADNP and antibody dependent complement deposition (ADCD). * p ≤ 0.05; ** p ≤ 0.01; *** p ≤ 0.001; **** p ≤ 0.0001.

Because Fc domain engagement is only the first step in signaling and initiation of effector functions, we examined the downstream consequences of activation by measuring RBD antibody dependent natural killer cell activation (ADNKA) which leads to antibody dependent cellular cytotoxicity (ADCC) (Chung et al., 2015), antibody dependent cellular and neutrophil phagocytosis (ADCP and ADNP) and antibody dependent complement deposition (ADCD). We found that neutralization titers positively correlated with all three markers of ADNKA: CD107a degranulation and intracellular IFNγ and TNFα production (Figure 2D-F). This association was not observed with ADNP (Figure 2G) and ADCP (Supplemental Figure 2A) and was less statistically significant with C3 deposition in ADCD (Supplemental Figure 2A). Because the primary Fc receptor that induces ADNKA is FcγRIIIa/CD16a, these findings corroborated data with respect to binding (Figure 2A). In contrast, the combinatorial engagement of low and high affinity FcγRs and the FcαR on neutrophils in ADNP did not correlate with neutralization (Figure 2G). Along these lines, the ratio of activating FcγRIIa/CD32a and, to a lesser degree, FcγRIIIa/CD16a, to the inhibitory FcγRIIb/CD32b involved in ADCP in THP-1 monocytes did not relate to neutralization (Supplemental Figure 2A and Figure 2H). The link between FcγRIIIa/CD16a NK cell activation and neutralization was sustained across variants, though fits again varied (Figure 2H and Supplemental Figure 2B). These data together demonstrated that in contrast to FcγRIIa/CD32a and FcγRIIb/CD32b, vaccine specific IgG induction of FcγRIIIa/CD16a functions associated with neutralization.

### IgG glycosylation

As in many infectious and non-infectious processes, post-translational glycosylation of polyclonal IgG has been shown to mediate binding affinity to Fc receptors in SARS-CoV-2 infection (Chakraborty et al., 2021; Chakraborty et al., 2022; Hoepel et al., 2021; Larsen et al., 2021). A core biantennary structure on the conserved asparagine residue N297 on the Fc domain is modified by the addition and subtraction of galactose (G), sialic acid (S), fucose (F) and bisecting N-acetylglucosamine (GlcNAc) to generate glycoform diversity (Arnold et al., 2007) (Supplemental Figure 3A). Monoclonal and polyclonal antibody studies have shown that changes in glycoform composition have the potential to impact binding and downstream effector functions (Supplemental Figure 3A) (Alter et al., 2018; Arnold et al., 2007; Peschke et al., 2017; Quast et al., 2015; van Osch et al., 2021). To evaluate the impact of glycosylation on vaccine induced antibodies, we measured the relative abundance of N-glycans on total non-antigen and RBD specific IgG (Supplemental Figure 3B). For each individual, non-antigen compared to RBD specific IgG glycoforms were distinct (Figure 3A and 3B). Glycoforms (Supplemental Figure 3C) containing fucose (Figure 3C), total sialic (Figure 3D) composed of di-sialic (Figure 3E) and mono-sialic (Figure 3F) acids, galactose (di-galactosylated in Figure 3G and agalactosylated and mono-galactosylated in Supplemental Figure 3D) and bisecting GlcNAc (Supplemental Figure 3E) were significantly different between total non-antigen and RBD specific IgG.

**Figure 3:**
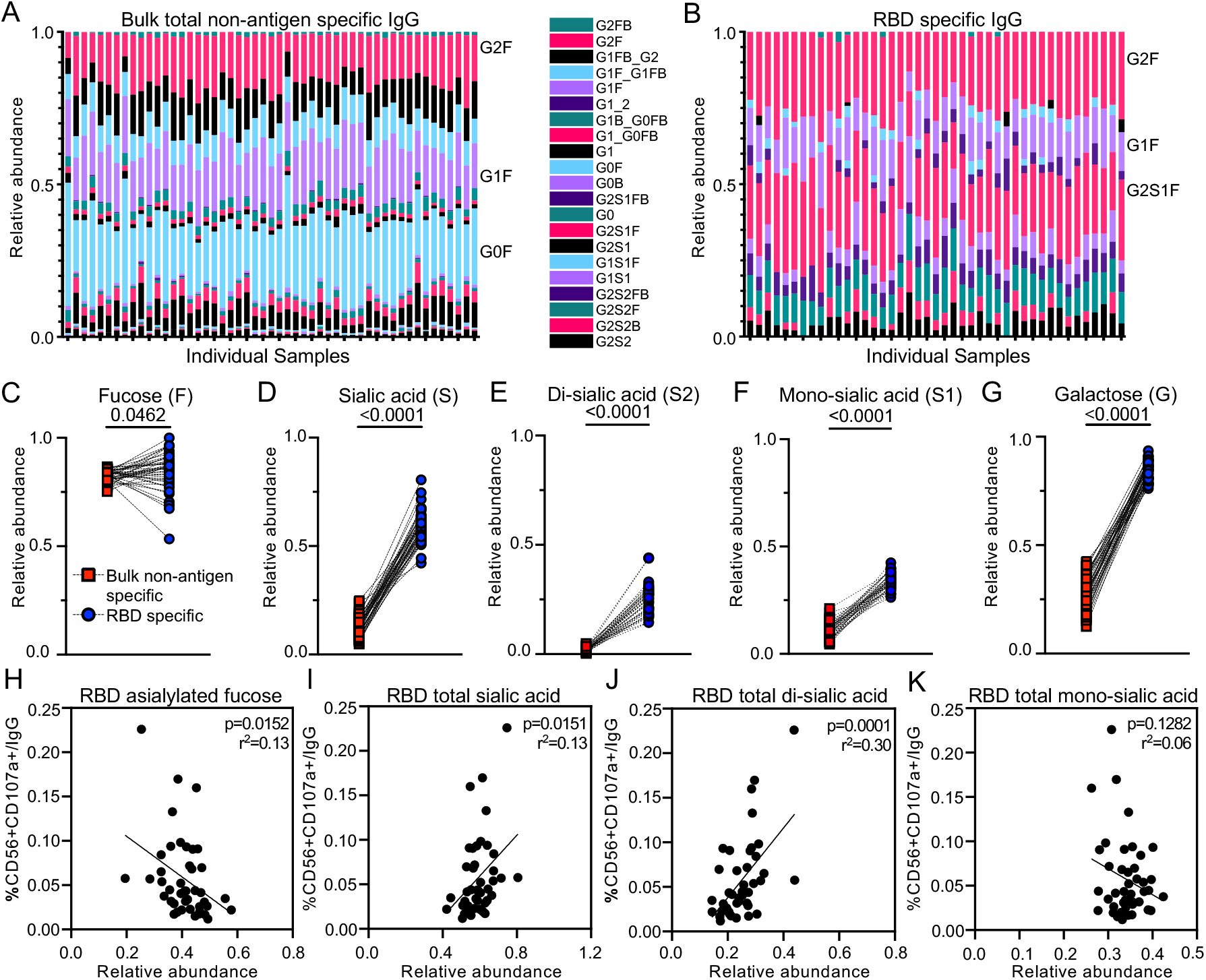
Differential fucose and sialic acid on vaccine specific IgG link to FcγRIIIa/CD16a effector functions. Stacked column graphs depict the relative abundance of individual glycoforms (Supplemental Figure 3A-B) with respect to (A) total bulk non-antigen specific and (B) receptor binding domain (RBD) specific IgG. Each column represents one individual study participant. Dot plots summarize differences between bulk non-antigen specific and RBD specific IgG in the collective relative abundance of all individual glycoforms (Supplemental Figure 3C and D) containing (C) fucose, (D) total sialic acid, (E) di-sialic acid, (F) mono-sialic acid and (G) di-galactose with statistical significance calculated by Wilcoxon matched-pairs signed rank test. Data for (H) asialylated fucosylated, (I) total sialic and (J) total di-sialic acid, the three RBD specific glycoforms that have a statistically significant relationship across all markers of ADNKA activation, are plotted with CD107a expression per RBD specific IgG, as well as IFNγ and TNFα (Supplemental Figure 4). For comparison, data for (K) total mono-sialic acid is plotted.

To evaluate if differential antibody glycosylation impacted effector functions associated with neutralization, we investigated which glycoforms lead to FcγRIIIa/CD16a mediated NK cell activation by linear regression. We found that relative levels of RBD and not total non-antigen specific IgG glycoforms significantly correlated with CD107a degranulation, intracellular IFNγ and TNFα production at varying levels (Supplemental Figure 4A-D). Relative levels of IgG glycoforms that contained fucose without sialic acid (asialylated fucosylated) and glycoforms that contained sialic acid (specifically di-sialic and not mono-sialic acid) correlated with all three markers of NK cell activation (Figure 3H-K). The negative relationship between asialylated fucosylated species on RBD specific IgG with ADNKA indicated an inhibitory effect of the presence of fucose. This contrasted with sialic acid, where the absence negatively (Figure 3H and Supplemental Figure 4E and H) and presence positively (Figure 3I and J and Supplemental Figure 4 F-G, I-J) associated with ADNKA. Taken together, these data showed that fucose and sialic acid on vaccine specific IgG influence FcγRIIIa/CD16a NK cell activation in opposing manners.

### Impact of age on antibody Fc effector functions

We next investigated if Fc domain features were dependent on age as we had observed with Fab domain mediated neutralization. We observed a negative relationship between RBD specific IgG binding to FcγRIIIa/CD16a, FcγRIIa/CD32a and FcγRIIb/CD32b with age (Figure 4 A-C) by linear regression taking sex into account (Figure 4D). In contrast, no statistically significant relationships between age and Fc receptor binding to antibodies targeting control antigens from other pulmonary viruses respiratory syncytial virus (RSV) and influenza (Flu) and the negative control *Bacillus anthracis* (Anthrax) were seen (Figure 4D). Consistent with neutralization data, we observed that age negatively correlated with RBD specific IgG mediated NK cell CD107a degranulation (Figure 4E) and intracellular IFNγ and TNFα production at varying levels (Supplemental Figure 5 and Figure 4F). In comparison, the relationships between age and RBD specific ADCP (Figure 4G), ADNP (Figure 4H) as well as ADCD (Supplemental Figure 5) were non-significant. Consistent with differential IgG glycosylation linked to NK cell activation (Figure 3), asialylated fuosylated glycoforms in RBD compared to non-antigen specific IgG were increased in those ≥65 years old (Figure 4I). These data showed that age negatively impacted some but not all Fc effector functions as it did for neutralization, which is likely due to the combination of decreased antibody levels and reduced antibody quality in differential glycosylation and altered FcR engagement.

**Figure 4:**
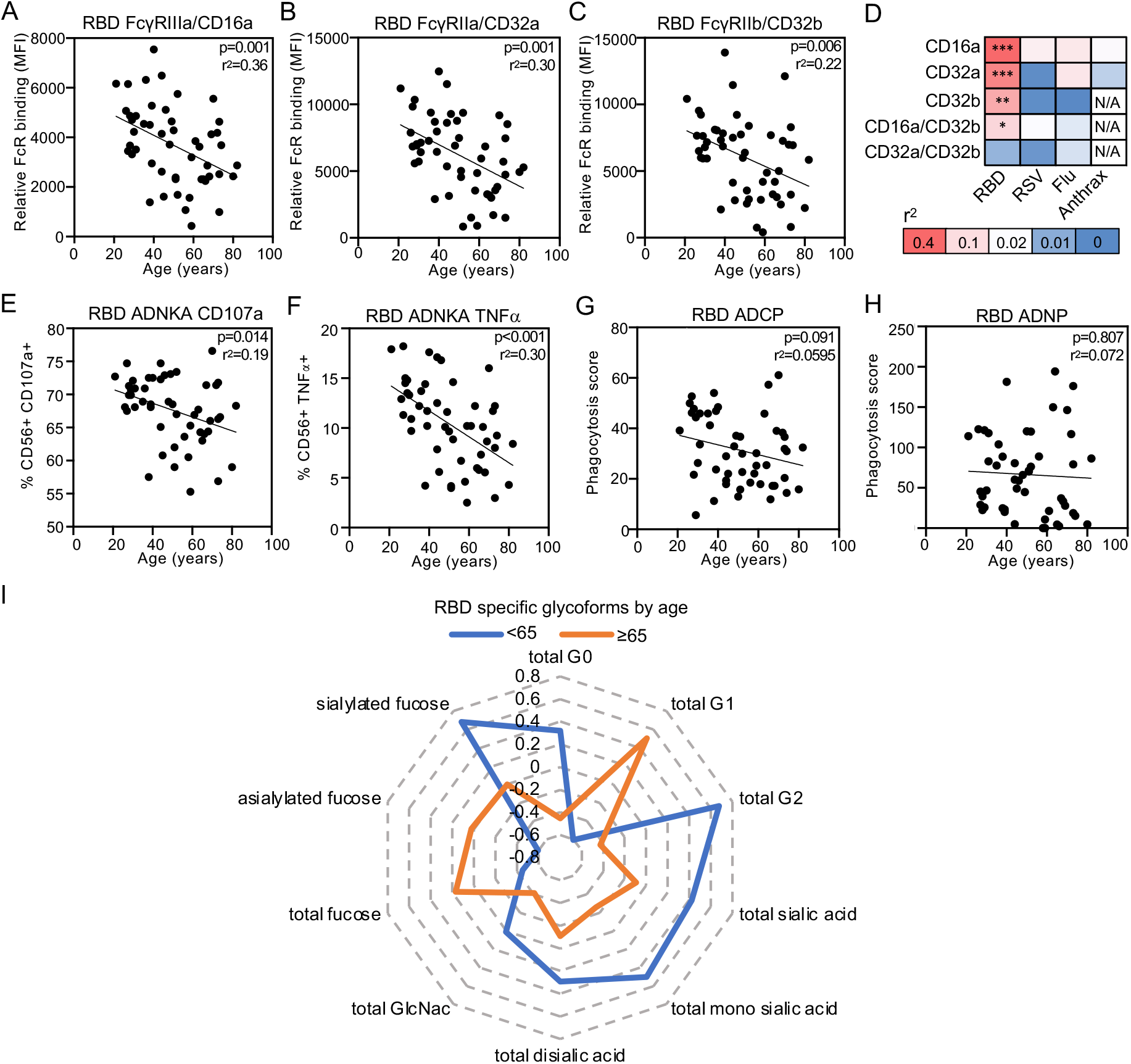
Age influences some but not all vaccine specific antibody FcγR functions. The relationships between relative binding of receptor binding domain (RBD) specific IgG to (A) FcγRIIIa/CD16a, (B) FcγRIIa/CD32a and (C) inhibitory FcγRIIb/CD32b and age are shown. (D) Heatmap of the coefficient of determination (r2) summarizes the goodness of fit across RBD and control respiratory syncytial virus (RSV), influenza (Flu) and anthrax antigens in FcγR binding and age. * p ≤ 0.05; ** p ≤ 0.01; *** p ≤ 0.001; N/A not available given absence of significant detectable levels. The relationship between age and RBD antibody dependent natural killer cell activation (ADNKA) as measured by (E) CD107a and (F) TNFα, and (G) RBD antibody dependent cellular phagocytosis (ADCP) and (H) RBD antibody dependent neutrophil phagocytosis (ADNP) are shown. Linear regression with p values adjusted for sex are reported. (I) Radar plots depict vaccine specific IgG glycoforms calculated from the Z scored data for each individual RBD specific IgG glycoforms relative to bulk non-antigen specific IgG glycoforms with lines representing the median for each age group.

### Polyclonal functional breadth and magnitude

Polyclonal antibody responses consist of multiple Fab and Fc domain features that interact to influence disease outcomes. To begin to assess the collective functionality for each vaccinee sample, we calculated the breadth of neutralization across all five SARS-CoV-2 isolates tested (Supplemental Figure 5A). In addition, we Z score transformed data from Fc assays to enable comparisons between effector functions and summarization of the cumulative Fc functional magnitude for each individual (Supplemental Figure 5B). To assess how Fc functionality related to Fab activity, we grouped individuals by their neutralization breadth. We found that neutralization of all variants (100%) was detectable in 28 of the 51 individuals, and those remaining demonstrated 50-83% breadth (Figure 5A). Of those with <100% neutralization breadth, the cumulative Fc functional scores were low or negative. Of those with 100% neutralization breadth, both positive and negative cumulative Fc functional scores were detected. Thus, high neutralization breadth and potent Fc effector functions are linked. Moreover, Fc functionality represented a source of immune variation in the presence of broad Fab mediated neutralization.

**Figure 5:**
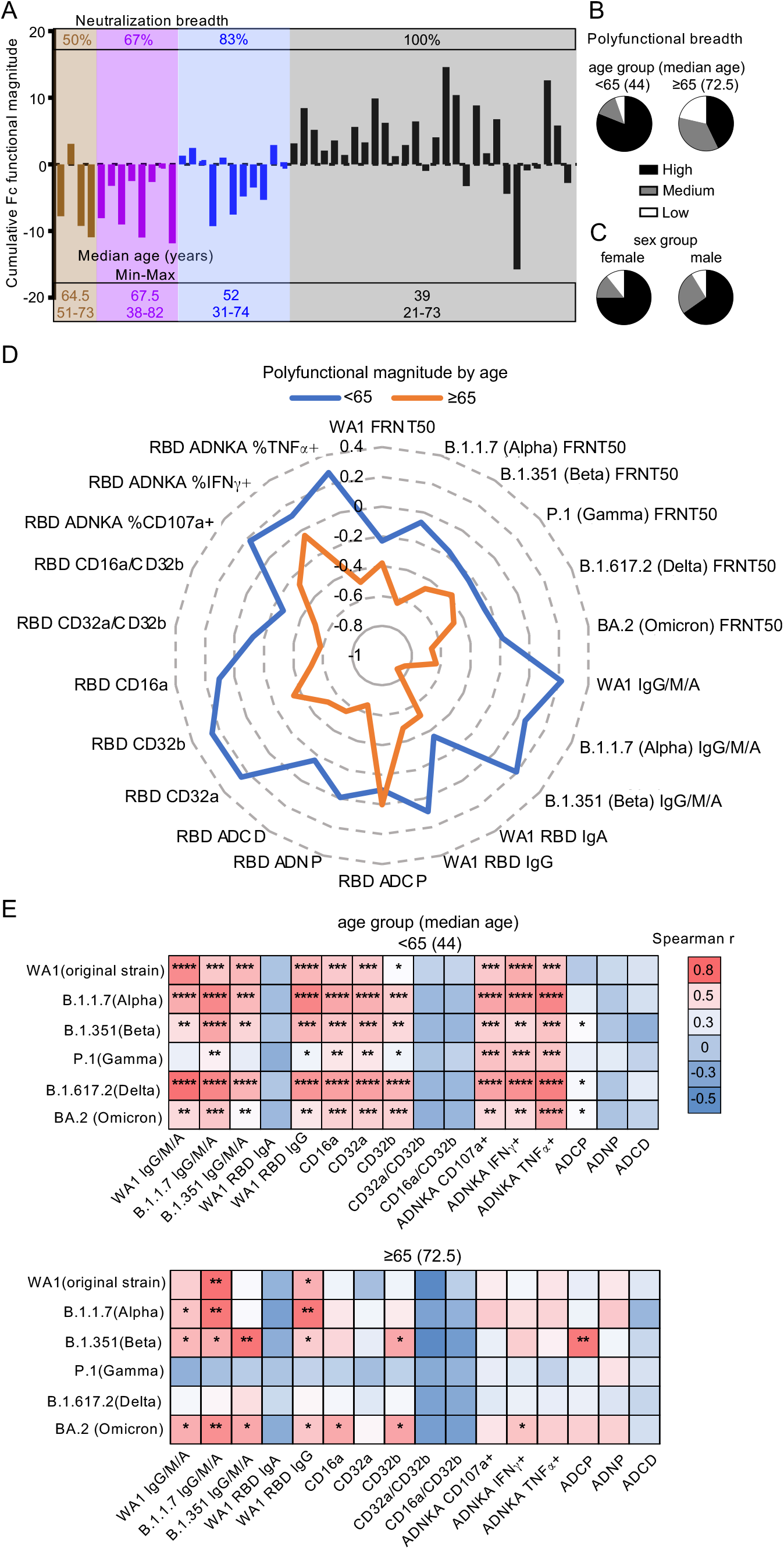
Enhanced BNT162b2 induced polyfunctional antibody breadth and magnitude against SARS-CoV-2 in younger compared to older adults. For each individual, the neutralization breadth across variants (Supplemental Figure 6A) and cumulative vaccine specific Fc functional magnitude from the sum of the Z scores for each of the individual effector functions (Supplemental Figure 6B) were calculated. (A) Grouped by neutralization breadth (top), each column shows the cumulative Fc functional score for one individual. Median, minimum and maximum ages characterizing each neutralization breadth group are shown (bottom). Polyfunctional antibody breadth was calculated for each individual (Supplemental Figure 6C) and used to categorize individuals into high (90-100%), medium (80-90%) or low (<80%) responders. The proportions of high, median and low responders are grouped by (B) age and (C) sex. (D) Radar plots depict vaccine specific polyfunctional antibody magnitude calculated from the Z scored data for each antibody function (Supplemental Figure 6C) with lines representing the median for each age group. (E) Heatmap summarizes Spearman correlations (Supplemental Figure 2) between neutralization of SARS-CoV-2 wildtype WA1 and variants with RBD specific IgG/M/A, IgG and IgA levels, relative binding of RBD specific IgG to activating (FcγRIIIa/CD16a and FcγRIIa/CD32a), inhibitory (FcγRIIb/CD32b) and ratios of activating:inhibitory FcγR (FcγRIIIa/CD16a:FcγRIIb/CD32b and FcγRIIa/CD32a:FcγRIIb/CD32b) binding and Fc effector functions antibody dependent natural killer cell activation (ADNKA), antibody dependent cellular phagocytosis (ADCP), antibody dependent neutrophil phagocytosis (ADNP) and antibody dependent complement deposition (ADCD) for each age group. * p ≤ 0.05; ** p ≤ 0.01; *** p ≤ 0.001; **** p ≤ 0.0001.

Because we observed that both neutralizing (Figure 1) and non-neutralizing (Figure 4) antibody functions were dependent on age, we assessed age with respect to neutralization breadth. We found that the median age for those with 100% neutralization was younger (39 years) compared to those with <100% (64.5, 67.5 and 52 years for 50%, 67% and 83% neutralization, respectively) (Figure 5A). Thus, both antibody Fab and Fc mediated breadth and potency diminished with age.

To focus on age categorically, we grouped individuals into those <65 and ≥65. The cutoff of 65 years was chosen for three reasons: 1) 63.5 is the median age of the subgroup of individuals with no detectable neutralization against Omicron (BA.2), the variant with the lowest overall activities (Figure 1B), 2) the median ages of the two groups with the lowest neutralization breadths are 64.5 and 67.5 (Figure 5A) and 3) ≥65 is the definition of older adults used by the Center for Disease Control and Prevention with respect to COVID-19 vaccine administration guidelines (Bialek et al., 2020). We calculated the polyfunctional breadth for each vaccinee by enumerating the proportion of detectable SARS-CoV-2 neutralizing and non-neutralizing responses to categorize individuals as high, medium, and low responders (Supplemental Figure 5C). We observed that most individuals <65 had high polyfunctional breadth while those ≥65 had low or medium (Figure 5B). This difference in breadth was not noted with groupings by sex (Figure 5C). In addition to antibody breadth, we evaluated polyfunctional magnitude using vaccine specific neutralizing and non-neutralizing antibody Z score data. We found that the extent of all antibody functions except for ADCP was diminished in the ≥65 compared to <65 group (Figure 5D). Because polyfunctional antibody responses are comprised of multiple activities that potentially occur concurrently to influence outcomes of infection, we assessed the coordination between antibody features and functions in these two age groups. We found more coordination in those <65 compared to ≥65 (Figure 5E). Thus, the breadth, magnitude, and coordination of BNT162b2 induced neutralizing and non-neutralizing antibody polyfunctionality diverge with respect to the age of 65 years.

## Discussion

In this study we show that two doses of the BNT162b2 mRNA vaccine elicited coordinated neutralizing and non-neutralizing antibody functions. The presence of vaccine specific antibodies is critical but neutralizing and non-neutralizing antibody functions are driven by quality as well as quantity. Thus, titers correlated with neutralizing activity (Figure 1A) and vaccine induced neutralizing responses against live clinical isolates of SARS-CoV-2 and five distinct variants decreased with age (Figure 1B). Neutralization correlated with FcγRIIIa/CD16a activation of natural killer cells that leads to cellular cytotoxicity but not phagocytosis or complement deposition (Figure 2). Engagement with FcγRIIIa/CD16a was associated with post-translational vaccine specific IgG afucosylation and sialylation (Figure 3H-K) which diverge with age (Figure 4I). Antibody functions were diminished among those aged ≥65: neutralization breadth across variants, overall Fc functional potency, and coordination between neutralizing and non-neutralizing antibody activities (Figure 5B, D, E), demonstrating compromised vaccine-induced polyfunctionality. Neutralizing activity and antibody titers are measured in vaccine studies to gauge effectiveness at blocking infection. The findings from this study show that non-neutralizing antibody effector functions are immune correlates that could inform on the potential of vaccines to prevent disease, a target which is of growing importance with the continual emergence of new variants that subvert neutralization.

Non-neutralizing antibody functions are mediated by immune complexing and binding between the Fc domain and Fc receptors. Thus, even with reduced Fab domain avidity for mutated viral proteins such as spike, vaccine induced non-neutralizing Fc functions could remain robust. Our data show that neutralizing activities across all variants are lower compared to wildtype virus (Figure 1B), suggesting that effectiveness in preventing infection is significantly compromised. However, even with increased case numbers of infection due to variants, epidemiological data show relatively strong vaccine protection against disease and hospitalization (Altarawneh et al., 2022; Andrews et al., 2022; Collie et al., 2022; Nasreen et al., 2022; Tang et al., 2021). Our data show that the correlation between titers and neutralizing activities was decreased across different variants, and the relationships with non-neutralizing functions, specifically ADNKA partially overlapped (Figure 5E).

In line with these observations from human studies, data from animal models demonstrate that *in vitro* neutralization does not uniformly correlate with *in vivo* protection against disease (Schafer et al., 2021). Moreover, enhancement of non-neutralizing Fc effector functions delay viral spread synergistically with neutralizing activity in mice (Beaudoin-Bussieres et al., 2022). In humans, our results show that many non-neutralizing Fc effector functions were elicited by vaccination but antibody dependent NK cell activation that leads to cellular cytotoxicity specifically linked to neutralization across wildtype and SARS-CoV-2 variants (Figure 2). Thus, along with inhibiting viral entry by neutralization, vaccine specific antibodies via FcγRIIIa/CD16a expressing NK cells, monocytes and macrophages could target cytotoxicity against airway epithelial cells already infected with SARS-CoV-2 to prevent viral spread and disease.

In natural infection, FcγRIIIa/CD16a is associated with disease severity (Chakraborty et al., 2021; Chakraborty et al., 2022; Hoepel et al., 2021; Junqueira et al., 2022; Larsen et al., 2021). While our data here do not include individuals with severe COVID-19 disease, the nature of polyclonal antibodies generated during natural infection diverge from vaccination. First, the antigenic repertoire after natural infection likely contains non-RBD specific antibody responses that are absent after vaccination. Second, antibody titers are likely diminished with exposure to lower amounts of antigen from mild and asymptomatic infection compared to severe disease and vaccination (Dufloo et al., 2021). Thus, FcγRIIIa/CD16a activities from immunity generated after natural infection could confer different downstream consequences compared to vaccination.

Post-translational IgG glycosylation influences Fc receptor binding and activation. Along with afucosylation that enhances FcγRIIIa/CD16a engagement which is also observed in severe COVID-19 disease, our data from whole vaccine specific IgG show that sialic acid could also contribute (Figure 3H and Figure 5). As such, sialylation on vaccine specific IgG could further modify FcγRIIIa/CD16a activation. The study of IgG glycosylation has focused primarily on N297 of the Fc domain (Chakraborty et al., 2021; Chakraborty et al., 2022; Farkash et al., 2021; Hoepel et al., 2021; Larsen et al., 2021), not accounting for the 20% of polyclonal IgG modified on the Fab domain (van de Bovenkamp et al., 2016). Our evaluation of whole IgG suggests that glycans from both Fab and Fc domains contribute to Fc effector functions by indirectly and directly affecting Fc receptor interactions (He et al., 2016; Shi et al., 2019; Yamaguchi et al., 2022). Thus, how an Fc receptor is activated by differential antibody glycosylation could be critical in determining the outcomes of downstream immune responses.

The factors which predict vaccine response at an individual level are the subject of intense study. Several lines of evidence support that age is one important factor (Bates et al., 2021a; Collier et al., 2021; Farkash et al., 2021). Our study was designed to look specifically at the contribution of age to non-neutralizing antibody activities from vaccination. In the elderly compared to younger individuals, virus specific memory B cells and antibody titers persist longer than neutralizing activity (Jeffery-Smith et al., 2022). Thus, loss of neutralization with a shift towards more dependence on non-neutralizing antibody activity could be a hallmark of immunosenescence. As such, monitoring non-neutralizing in addition to neutralizing functions could help determine the need and dose of booster vaccinations for this population. Moreover, approaches using adjuvants to enhance vaccine mediated non-neutralizing antibody functions such as FcγRIIIa/CD16a could be beneficial (Coler et al., 2018).

With respect to the broader population, analyses of longitudinal and cross-sectional studies involving vaccination and infection show that non-neutralizing functions including NK cell activity and ADCC are sustained longer than neutralization (Fuentes-Villalobos et al., 2022; Lee et al., 2021; Tso et al., 2021). Modeling of neutralization decay predicts that protection from infection is lost but protection from severe disease is retained (Khoury et al., 2021). This divergence between neutralizing titers and immune protection is likely due to multiple factors including viral fitness (Mlcochova et al., 2021; Wang et al., 2021; Weisblum et al., 2020), T cell activities (Keeton et al., 2022) as well as non-neutralizing responses such as the FcγRIIIa/CD16a functions observed here. Thus, enhancing non-neutralizing activities elicited by vaccines could provide longer lasting protection against disease independent of altering vaccine antigens to target each new variant.

Current CDC vaccine recommendations for healthy adults <50 involve three total doses and for those ≥50, four. At the time of this writing, 91.8% of the US population ≥65 who have received two doses, 70.4% three and 39.1% four (CDC, 2022). Outside the US, many parts of the world still have limited access to vaccine and have lower rates of vaccination. Our data support the assertion that for those elderly individuals with two doses of BNT162b2, immunity is suboptimal because neutralizing and non-neutralizing antibody activities are restricted. The effects of additional doses of vaccines using antigens from the original SARS-CoV-2 strain or Omicron and infection on top of vaccination that generates hybrid immunity remain to be fully defined but likely encompass enriched neutralization breadth and Fc potency (Collier et al., 2021; Farkash et al., 2021; Richardson et al., 2022). How much protection is enhanced is a subject of active discourse (Atmar et al., 2022; Regev-Yochay et al., 2022). Evaluating the breadth, magnitude and coordination of polyclonal antibody functions (Figure 5) will enhance resolution of correlates of protection, particularly in the context of variants where the effect of neutralizing activity is likely limited. There is growing evidence that adjuvants and antigens can be used to skew immune responses including antibody glycosylation and Fc effector functions for rational vaccine design (Bartsch et al., 2020; Boudreau et al., 2020; Mahan et al., 2016; Oefner et al., 2012). Approaches that leverage the collaboration between antibody Fab and Fc domain functions could improve vaccine efficacy against variants for all, and specifically for vulnerable populations with difficulty generating neutralizing responses such as the elderly.

### Limitations of the study

Limitations to this study include sample size, the lack of ethnicity, race and clinical data and the homogeneity of the population examined with all participants being employees of a local health care system. These cohort characteristics limited the ability to resolve more subtle differences and extrapolate across a diverse array of individuals but also minimized potential sources of confounding variables, likely facilitating the discovery of relationships between antibody features that would otherwise be difficult to discern due to the complexity and heterogeneity of polyfunctional antibodies. The absence of infection was not determined by molecular microbiological diagnostics but rather serologically by the lack of detectable nucleocapsid (Supplemental Figure 1A), RBD specific antibodies prior to vaccination (Bates et al., 2021a) and clinical history. As such, it is plausible that individuals with asymptomatic infections are included. However, the dominant immune responses measured were likely due to vaccination given the narrow window between the second vaccine dose and sample collection time (14-15 days). As the cohort was sex balanced, the major known phenotypic variation in this group was age (21 to 82 years).

## STAR Methods

### RESOURCE AVAILABILITY

#### Lead contact

Further information and requests for resources and reagents should be directed to and will be fulfilled by the lead contact, Lenette Lu.

#### Materials availability

No unique reagents were generated during the course of this study.

#### Data and code availability

The dataset generated during this study is available upon reasonable request. This paper does not report original code. Any additional information required to reanalyze the data reported in this paper is available from the lead contact upon request.

### EXPERIMENTAL MODEL AND SUBJECT DETAILS

#### Cohort

Study participants (n=51) were enrolled between December 2020 and February 2021 at Oregon Health & Science University immediately after receiving their first dose of BNT162b2 vaccine. Participants received a second vaccine dose between 21±1 days following the first dose, then returned 14-15 days later for follow up. Whole blood was collected in serum tubes (BD) and serum isolated by centrifugation 1000xg for 10min. Sera were heat inactivated at 65°C for 30min then frozen at -20°C. This study was conducted in accordance with the Oregon Health & Science University Institutional Review Board with written informed consent from all participants, and approved by the UT Southwestern Medical Center Institutional Review Board. Written informed consent was received from all study participants prior to participation.

#### Cell Lines

Vero E6 cells were purchased from ATCC (ATCC VERO C1008), grown at 37C, 5% CO2 and maintained in Dulbecco’s Modified Eagle Medium supplemented with 10% fetal bovine serum, 1% penicillin/streptomycin, 1% non-essential amino acids. THP-1 cells were purchased from ATCC (ATCC TIB-202), grown at 37C, 5% CO2 and maintained in RPMI-1640 supplemented with 10% fetal bovine serum, 2mM L-glutamine, 10mM HEPES, and 0.05 mM β-mercaptoethanol. CD16.NK-92 (ATCC PTA-6967) were purchased from ATCC (ATCC PTA-6967), grown at 37C, 5% CO2 and maintained in in MEM-α supplemented with 12.5% FBS, 12.5% horse serum, 1.5g/L sodium bicarbonate, 0.02mM folic acid, 0.2mM inositol, 0.1 mM 2-β-mercaptoethanol, 100U/mL recombinant IL-2.

#### Primary Immune Cells

Fresh peripheral blood was collected at UT Southwestern from healthy volunteers. All were over 18 and de-identified prior to blood processing. Neutrophils isolated from peripheral blood were maintained at 37C, 5% CO2 in RPMI with 10% fetal bovine serum, L-glutamine, and HEPES. The study was approved by the UT Southwestern Medical Center Institutional Review Board. Written informed consent was received from all study participants prior to participation.

### METHOD DETAILS

#### Virus

SARS-CoV-2 clinical isolates were passaged once before use in neutralization assays: USA-WA1/2020 [original strain] (BEI Resources NR-52281); USA/CA_CDC_5574/2020 [B.1.1.7] (BEI Resources NR-54011); hCoV-54 19/South Africa/KRISP-K005325/2020 [B.1.351] (BEI Resources NR-54009); hCoV-19/Japan/TY7-503/2021 [P.1] (BEI Resources NR-54982); hCoV-19/USA/PHC658/2021 [B.1.617.2] (BEI Resources NR-55611); and hCoV-19/USA/CO-CDPHE-2102544747/2021 [B.1.1.529 - BA.2] (BEI Resources NR-56520). Isolates were propagated in Vero E6 cells for 24 to 72hrs until cultures displayed at least 20% cytopathic effect (CPE), as previously described.

### Enzyme Linked Immunosorbent Assays (ELISA)

ELISAs were performed as described (Bates et al., 2021b). Plates were coated overnight at 4°C with 1 mg/mL recombinant SARS-CoV-2 spike receptor binding domain (RBD) protein (Bates et al., 2021c) (BEI Resources NR-52309) or recombinant SARS-CoV-2 nucleocapsid (N) protein (BEI Resources NR-53797). Serum dilutions (6 × 3-fold for RBD, 6 × 4-fold for N) in duplicate were prepared in 5% milk powder, 0.05% Tween-20, in phosphate buffered saline (PBS), starting at 1:1600 (pan-Ig), 1:50 (IgA), 1:200 (IgG). The secondary antibodies used were pan-Ig (1:10,000 anti-human GOXHU IgG/A/M-HRP, A18847 Invitrogen), IgA (1:3,000 anti-human IgA-HRP, 411002 Biolegend), and IgG (1:3,000 anti-human IgG-HRP 555788, BD Biosciences). Plates were developed with o-phenylenediamine (OPD) (ThermoScientific). Absorbance at 492nm was measured on a CLARIOstar plate reader and normalized by subtracting the average of negative control wells and dividing by the highest concentration from a positive control dilution series. ELISA EC50 values were calculated by fitting normalized A492 as described (Bates et al., 2021b). The limit of detection (LOD) was defined by the lowest dilution tested for RBD and half of the lowest dilution for N. Values below the LOD were set to LOD – 1.

#### Focus Reduction Neutralization Test (FRNT)

Focus forming assays were performed as described (Bates et al., 2021b). Sub-confluent Vero E6 cells were incubated for 1 hour with 30 µL of diluted sera (5 × 4-fold starting at 1:20) which was pre-incubated for 1 hour with 100 infectious viral particles per well. Samples were tested in duplicate. Wells were covered with 150 µL of overlay media containing 1% methylcellulose and incubated for 24hrs, 48hrs for Omicron. Plates were fixed by soaking in 4% formaldehyde in PBS for 1 hour at room temperature. After permeabilization with 0.1% BSA, 0.1% saponin in PBS, plates were incubated overnight at 4°C with primary antibody (1:5,000 anti-SARS-CoV-2 alpaca serum, 1:2,000 for Omicron) (Capralogics Inc) (Bates et al., 2021b). Plates were then washed and incubated for 2hrs at room temperature with secondary antibody (1:20,000 anti-alpaca-HRP, 1:5,000 for Omicron) (NB7242 Novus) and developed with TrueBlue (SeraCare) for 30min. Foci were imaged with a CTL Immunospot Analyzer, enumerated using the viridot package (Katzelnick et al., 2018) and percent neutralization calculated relative to the average of virus-only wells for each plate. FRNT50 values were determined by fitting percent neutralization to a 3-parameter logistic model as described previously (Bates et al., 2021b). The limit of detection (LOD) was defined by the lowest dilution tested, values below the LOD were set to LOD – 1. Duplicate FRNT50 values were first calculated separately to confirm values were within 4-fold. When true, a final FRNT50 was calculated by fitting to combined replicates.

#### Fc receptor binding assays

Fc receptor binding assays were performed as described with modifications (Brown et al., 2017). Carboxylated microspheres (Luminex) were coupled with recombinant SARS-CoV-2 RBD (Bates et al., 2021c)(BEI Resources NR-52309) by covalent NHS-ester linkages via EDC (1-Ethyl-3-[3-dimethylaminopropyl]carbodiimide hydrochloride, Thermo Scientific Pierce) and Sulfo-NHS (N-hydroxysulfosuccinimide) (Thermo Scientific) per the manufacturer’s instructions. A mixture of influenza antigens from strain H1N1 (NR-20083 and NR-51702, BEI Resources), H5N1 (NR-12148, BEI Resources), H3N2, B Yamagata lineage, and B Victoria lineage (NR-51702, BEI Resources) was used as a control. A mixture of recombinant *Bacillus anthracis* antigens (Anthrax Protective Antigen, NR-36208 BEI Resources; Anthrax Lethal Factor, NR-28544 BEI Resources; Anthrax Edema Factor, NR-36210 BEI Resources) and a separate mixture of recombinant Respiratory Syncytial Virus antigens (G protein from strain B1, NR-31098 BEI Resources; F protein from strain B1, NR-31097 BEI Resources; G protein from strain A2, NR-31096 BEI Resources) were also used as controls. Antigen-coupled microspheres (1250 beads per well) were incubated with serially diluted sera (1:100, 1:1000, 1:10000) in 96-well Bioplex Pro Flat Bottom plates (Bio-Rad) at 4°C for 16hrs. Recombinant Fc receptors (FcγRIIIa/CD16a, FcγRIIa/CD32a, FcγRIIb/CD32b, R&D Systems) were fluorescently labeled with PE (Abcam) before addition to bead bound antigen specific immune complexes. After 2hrs of incubation at room temperature, the beads were washed with PBS with 0.05% Tween20 and antigen specific antibody bound Fc receptor measured on a on a MAGPIX instrument containing xPONENT4.2 software (Luminex). The background signal, defined as MFI of microspheres incubated with PBS, was subtracted. Representative data from one dilution was chosen by the highest signal to noise ratio for further analyses.

#### Non-antigen and RBD-specific IgG glycosylation

Non-antigen and RBD specific IgG glycans were purified and relative levels quantified as described with modifications (Mahan et al., 2015; Varadi et al., 2014). Recombinant RBD protein (BEI Resources NR-52309) (Bates et al., 2021c) was biotinylated with sulfosuccinimidyl-6-[biotinamido]-6-hexanamido hexanoate (sulfo-NHS-LC-LC biotin; ThermoScientific) and coupled to streptavidin beads (New England Biolabs). Patient sera were incubated with RBD-coupled beads and excess sera washed with PBS (Sigma). Bead-bound RBD-specific antibodies then eluted using 100mM citric acid (pH 3.0) and neutralized with 0.5M potassium phosphate (pH 9.0). Non-antigen specific or RBD-specific IgG were purified from the serum or eluted RBD-specific antibodies respectively by protein G beads (Millipore). Purified IgG was denatured and treated with PNGase enzyme (New England Biolabs) for 12hrs at 37°C to release glycans.

To isolate bulk IgG glycans, proteins were removed by precipitation using ice cold 100% ethanol at -20°C for 10min. To isolated RBD-specific IgG glycans, Agencourt CleanSEQ beads (Beckman Coulter) were used to bind glycans in 87.5% acetonitrile (Fisher Scientific). The supernatant was removed, glycans eluted from beads with HPLC grade water (Fisher Scientific) and dried by centrifugal force and vacuum (CentriVap). Glycans were fluorescently labeled with a 1.5:1 ratio of 50mM APTS (8-aminoinopyrene-1,3,6-trisulfonic acid, ThermoFisher) in 1.2M citric acid and 1M sodium cyanoborohydride in tetrahydrofuran (Fisher Scientific) at 55°C for 3hrs. The labeled glycans were dissolved in HPLC grade water (Fisher Scientific) and excess unbound APTS was removed using Agencourt CleanSEQ beads and Bio-Gel P-2 (Bio-rad) size exclusion resin. Glycan samples were run with a LIZ 600 DNA ladder in Hi-Di formamide (ThermoFisher) on an ABI 3500xL DNA sequencer and analyzed with GlycanAssure Data Acquisition Software v.1.0. Each glycoform was separated by peaks and identified based on glycan standard libraries (GKSP-520, Agilent). The relative abundance of each glycan for each individual sample was determined as (area under curve of each glycan)/ (sum of area under curve of all individual glycans).

#### Antibody dependent cellular phagocytosis (ADCP)

The THP-1 (TIB-202, ATCC) phagocytosis assay of antigen-coated beads was conducted as described with modifications (Lu et al., 2016). SARS-CoV-2 RBD recombinant protein (BEI Resources NR-52309) (Bates et al., 2021c) was biotinylated with Sulfo-NHS-LC Biotin (Thermo Fisher), then incubated with 1 μm fluorescent neutravidin beads (Invitrogen) at 4°C for 16hrs.

Excess antigen was washed away and RBD-coupled neutravidin beads were resuspended in PBS-0.1% bovine serum albumin (BSA). RBD-coupled beads were incubated with serial dilutions of sera (1:100, 1:500 and 1:2500) in duplicate for 2hrs at 37°C. THP1 cells (1×10^5^ per well) were then added. Plasma opsonized RBD-coupled beads and THP1 cells were incubated at 37°C for 16hrs. Cells were then washed once and fixed with 4% PFA. Bead uptake was measured on a BD LSRFortessa (SCC) equipped with high-throughput sampler and analyzed by FlowJo10. Phagocytic scores were calculated as the integrated median fluorescence intensity (MFI) (% bead-positive frequency × MFI/10,000) (Darrah et al., 2007). Representative data from one dilution was chosen by the highest signal to noise ratio for further analyses.

#### Antibody dependent neutrophil phagocytosis (ADNP)

The neutrophil phagocytosis assay of antigen-coated beads was conducted as described with modifications (Lu et al., 2016). Whole healthy donor blood was mixed with equal volume 3% dextran-500 (Thermo Fisher) and incubated for 25 min at room temperature to lyse and pellet the red blood cells. Leukocytes were removed and washed in endotoxin-free sterile water (Cytiva), followed by 1.8% NaCl (Thermo scientific) and then Hanks’ balanced salt solution without calcium and magnesium (Thermo Fisher). RBD conjugated beads, as described above, were incubated with serial dilution of sera (1:100, 1:500 and 1:2500) for 2hrs at 37°C. Isolated neutrophils (1 × 10^5^ per well) were added and incubated for 2hrs at 37°C. Bead uptake was measured on a BD LSRFortessa (SCC) equipped with high-throughput sampler and analyzed by FlowJo10. Phagocytic scores were calculated as the integrated median fluorescence intensity (MFI) (% bead-positive frequency × MFI/1,000). The purity of neutrophils was confirmed by staining with CD66b (BioLegend). Sera samples were tested in two independent experiments with neutrophils from two different HIV negative healthy donors. The mean of the data from both donors was used for further analysis. Representative data from one dilution was chosen by the highest signal to noise ratio for further analyses.

#### Antibody dependent complement deposition (ADCD)

The ADCD assay was performed as described with modifications (Fischinger et al., 2019). Carboxylated microspheres (Luminex) were coupled with SARS-CoV-2 RBD protein (Bates et al., 2021c) (NR-52309 BEI Resources) by covalent NHS-ester linkages via EDC (1-Ethyl-3-[3-dimethylaminopropyl]carbodiimide hydrochloride, Thermo Scientific Pierce) and Sulfo-NHS (N-hydroxysulfosuccinimide, Thermo Scientific) per manufacturer instructions. A mixture of influenza antigens from strains H1N1 (NR-20083 and NR-51702, BEI Resources), H5N1 (NR-12148, BEI Resources), H3N2, B Yamagata lineage, and B Victoria lineage (NR-51702, BEI Resources) was used as a control. Serum samples were heated at 56°C for 30min. Antigen-coated microspheres (1250 per well) were added to a 96-well Bioplex Pro Flat Bottom plates (Bio-Rad) and incubated with serial dilutions of sera (1:10, 1:50 and 1:250) in duplicate at 4°C for 16hrs. Freshly resuspended lyophilized guinea pig complement (Cedarlane) diluted 1:60 was added to the plate for 20min at 37°C. After washing off excess complement three times with15mM EDTA, anti-C3 PE-conjugated goat polyclonal IgG (MP Biomedicals) was added. The beads were then washed and C3 deposition quantified on a MAGPIX instrument containing xPONENT4.2 software (Luminex). The background signal, defined as MFI of microspheres incubated with PBS, was subtracted. Representative data from one dilution was chosen by the highest signal to noise ratio for further analyses.

#### Antibody dependent NK cell activation (ADNKA)

ADNKA assay was performed as described with modifications (Gunn et al., 2020). ELISA plates were coated with recombinant RBD antigen (300 ng/well) (Bates et al., 2021c) (BEI Resources NR-52309). Wells were washed, blocked, and incubated with serial dilutions of sera (1:10, 1:30, 1:90) in duplicate for 2hrs at 37°C prior to adding CD16a.NK-92 cells (PTA-6967, ATCC) (5 × 10^4^ cells/well) for 5hrs with brefeldin A (Biolegend), Golgi Stop (BD Biosciences) and anti-CD107a (clone H4A3, BD Biosciences). Cells were stained with anti-CD56 (clone 5.1H11, BD Biosciences) and anti-CD16 (clone 3G8, BD Biosciences) and fixed with 4% PFA. Intracellular cytokine staining to detect IFNγ (clone B27, BD Biosciences) and TNFα (clone Mab11, BD Biosciences) was performed in permeabilization buffer (Biolegend). Markers were measured using a BD LSRFortessa and analyzed by FlowJo10. CD16 expression was confirmed in all cells. NK cell degranulation and activation were calculated as percent of CD56+NK cells positive for CD107a, or IFNγ or TNFα expression. Representative data from one dilution was chosen by the highest signal to noise ratio for further analyses.

### QUANTIFICATION AND STATISTICAL ANALYSIS

Statistical analysis and graphing were performed using Stata17 and GraphPad Prism9.0. Data are summarized using the descriptive measures median, minimum, maximum and percent (%). Wilcoxon matched pair signed rank tests were used to compare neutralization of live SARS-CoV-2 variants (Figure 1B) and glycoforms between antigen non-specific and RBD specific IgG (Figures 3C-F, Supplemental Figure 3D-E). Mann-U-Whitney tests were used to compare the neutralization of live SARS-CoV-2 variants between male and female (Supplemental Figure 1C). Spearman rank correlations were used to examine bivariate associations between variables (Figure 2 and 5E, Supplemental Figure 1D and 2). Simple linear regression was used to examine the relationship between IgG glycoforms as the independent and Fc functional profiles as the dependent variables (Figure 3H-K and Supplemental Figure 4). Multiple robust regression models were used to adjust for the effect of age and sex when comparing the study variables between individuals (Figure 1A, C-H, Figure 4, and Supplemental Figure 5). Z scores of each individual Fc feature was calculated and then summed to generate the cumulative Fc functional magnitude (Figure 5A, Supplemental Figure 6B). For the radar plots (Figure 4I), Z scores of each individual RBD specific IgG glycoforms relative to bulk non-antigen specific IgG glycoforms were calculated and the median values for each age group were plotted. For the radar plots (Figure 5D), Z scores of each feature for each individual were calculated and the median values for each group were plotted. All p values are two-sided, and p < 0.05 was considered significant. In figures, asterisks denote statistical significance (* p ≤ 0.05; ** p ≤ 0.01; *** p ≤ 0.001; *** p ≤ 0.0001) with comparisons specified by connecting lines.

## Acknowledgements

This study was funded by a grant from the M. J. Murdock Charitable Trust (to MEC), an unrestricted grant from the OHSU Foundation (to MEC), the NIH training grant T32HL083808 (to TAB), NIH grant R011R01AI141549-01A1 (to FGT), OHSU Innovative IDEA grant 1018784 (to FGT), NIH grant R01AI145835 (to WBM), Burroughs Wellcome Fund UT Southwestern Training Resident Doctors as Innovators in Science (to YJK), pilot project grant from the UT Southwestern Department of Internal Medicine and Disease Oriented Scholars Award (to LLL). We gratefully acknowledge the OHSU workforce members who participated in this study; the OHSU COVID-19 serology study team and the OHSU occupational health department for their efforts in recruitment and sample acquisition; the OHSU clinical laboratory under the direction of Donna Hansel and Xuan Qin for SARS-Co-2 testing and reporting. We thank UTSW healthy volunteers who donated their blood for neutrophil studies; Dawn Wetzel for efforts in recruitment; Gabrielle Lessen for phlebotomy assistance; Ann McDonald for graphical assistance. We are grateful for the support of the M.J. Murdock Charitable Trust and the OHSU Foundation. The funders of the study had no role in study design, execution, analysis, interpretation, or writing of this manuscript.

## Author Contributions

LLL and FGT conceived, designed and supervised the work. TAB and PL designed, conducted and analyzed experiments. MEC, WBM, DS, SKM coordinated sample and reagent collection. SKM acquired and analyzed data. YJK, MT, CP, and DK analyzed the data. LLL, FGT, TAB, and PL wrote the manuscript. YJK, MT, SKM, DK, WBM, MEC contributed to manuscript revisions.

---

**Supplemental Figure 1:**
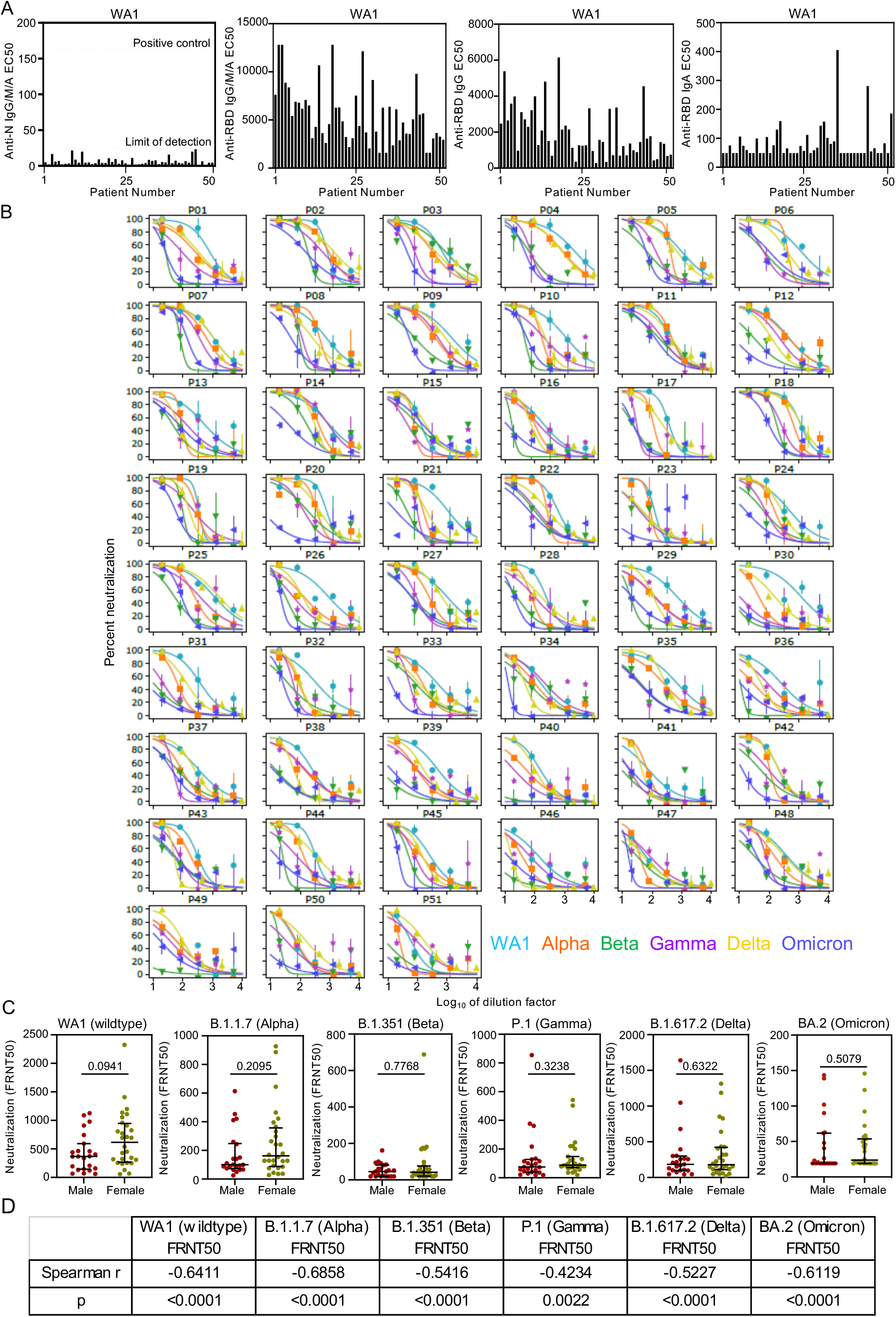
BNT162b2 vaccination induces IgG mediated neutralization of SARS-CoV-2 wildtype (WT) and clinical variants that diminish with age but are not altered by sex. (A) EC50 values are depicted for each individual for Nucleocapsid (N) specific antibodies, receptor binding domain (RBD) specific antibodies, IgG and IgA. Each column represents one individual. (B) Neutralization graphs from focus forming assays to calculate FRNT50 for each SARS-CoV-2 WT and clinical variants are shown. Each graph shows the data for one individual. (C) Dot plots show the distribution of neutralization for SARS-CoV-2 WT and variants by sex with statistical significance calculated by Mann-U-Whitney. (D) Spearman correlation coefficients and statistical significance between age and neutralization for SARS-CoV-2 WT and clinical variants are shown.

**Supplemental Figure 2:**
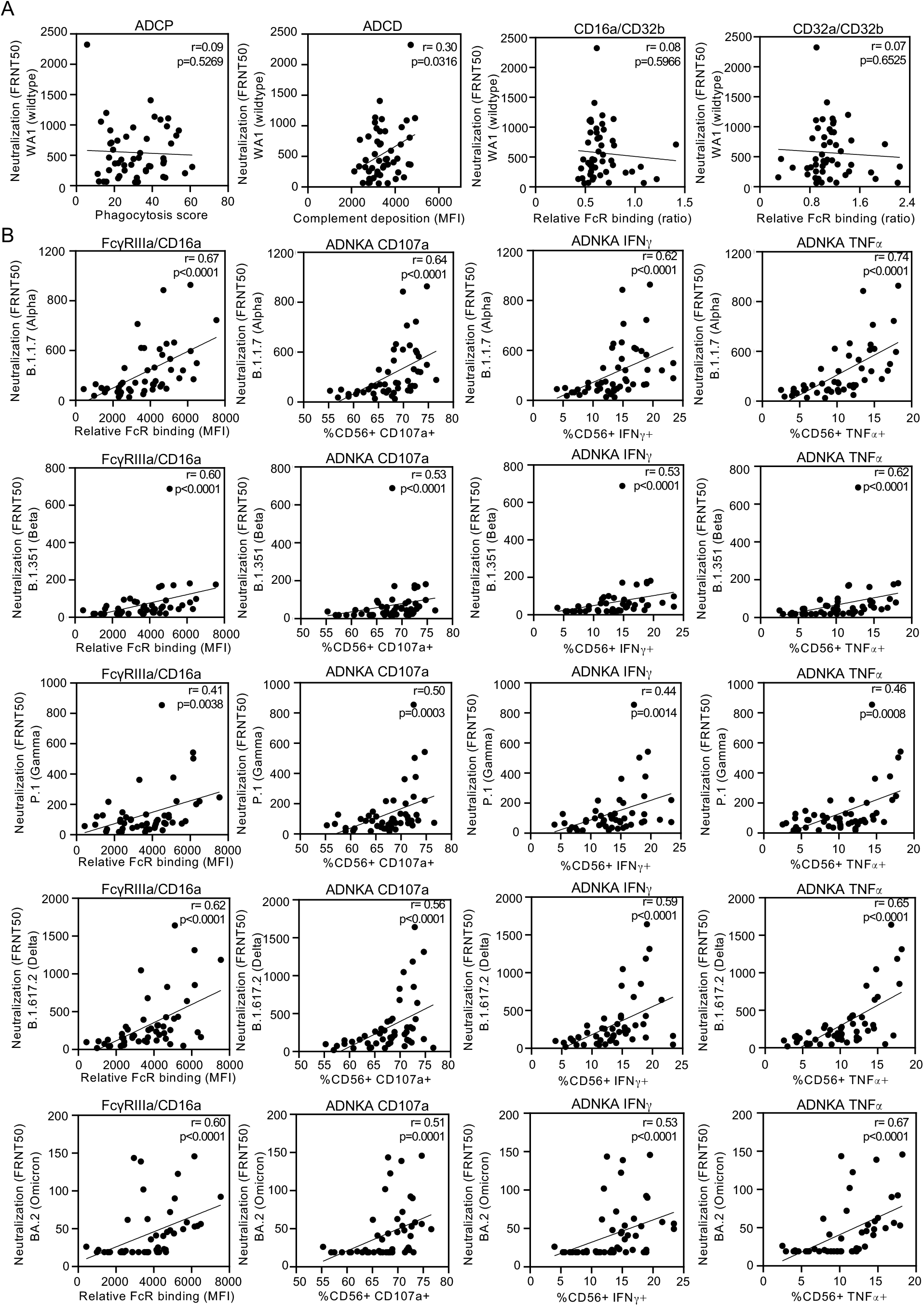
Vaccine specific IgG induction of FcγRIIIa/CD16 effector functions correlate with neutralization of SARS-CoV-2 variants. (A) The relationships between live SARS-CoV-2 WA1 wildtype neutralization and RBD specific antibody dependent cellular phagocytosis (ADCP), and RBD specific antibody dependent complement deposition (ADCD), and receptor binding domain (RBD) specific relative binding ratios of activating:inhibitory FcγR FcγRIIIa/CD16a:FcγRIIb/CD32b, FcγRIIa/CD32a:FcγRIIb/CD32b are shown. (B) The relationships between live SARS-CoV-2 variants neutralization and RBD specific FcγRIIIa/CD16a binding and effector function antibody dependent natural killer cell activation (ADNKA) are depicted. Statistical significances were determined by Spearman correlation.

**Supplemental Figure 3:**
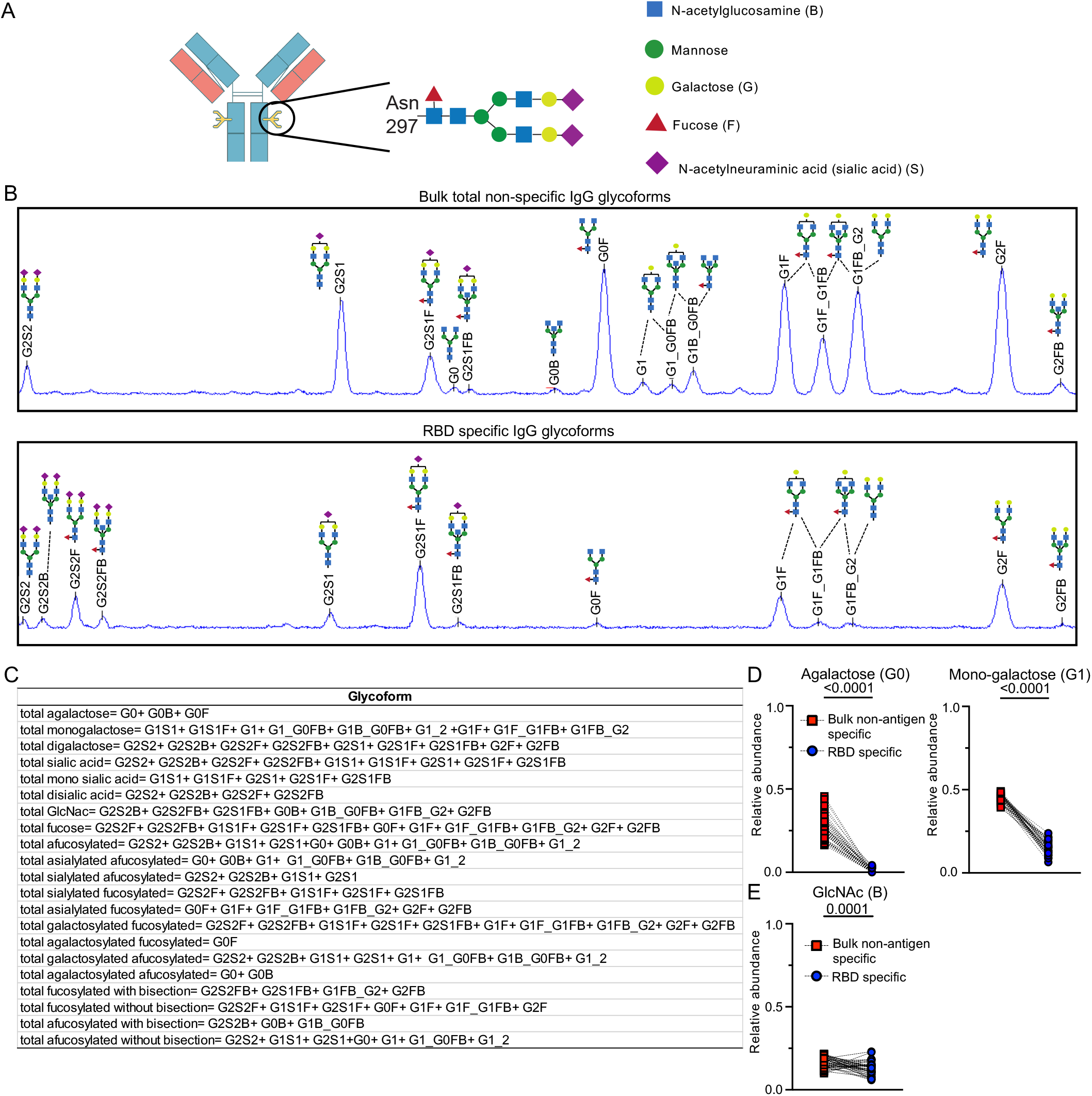
Bulk total non-antigen and vaccine specific IgG glycosylation patterns diverge. (A) Human IgG1 contains a conserved Fc domain N297 residue on which a bi-antennary structure of N-acetylglucosamine (GlcNAc) and mannose resides. The subsequent addition and subtraction of galactose (G), fucose (F), N-acetylneuraminic acid (sialic acid) (S) and bisecting GlcNAc (B) contributes to post translational diversity that develops with antibody maturation through the Golgi and ER. (B) Capillary electrophoresis chromatographs for bulk total non-antigen and receptor binding domain (RBD) specific IgG glycans captured from one individual are shown. Quantification of each peak determines the relative abundance of each glycoform depicted. (C) The collective relative abundance of all individual glycoforms with fucose (F), sialic acid (S), galactose (G) and bisecting GlcNAc (B) are calculated for bulk total non-antigen and RBD specific IgG. Differences between bulk total non-antigen and RBD specific (D) agalactosylated and mono-galactosylated and (E) bisecting GlcNAc structures are shown. Statistical significances were calculated by Wilcoxon matched-pairs signed rank test.

**Supplemental Figure 4:**
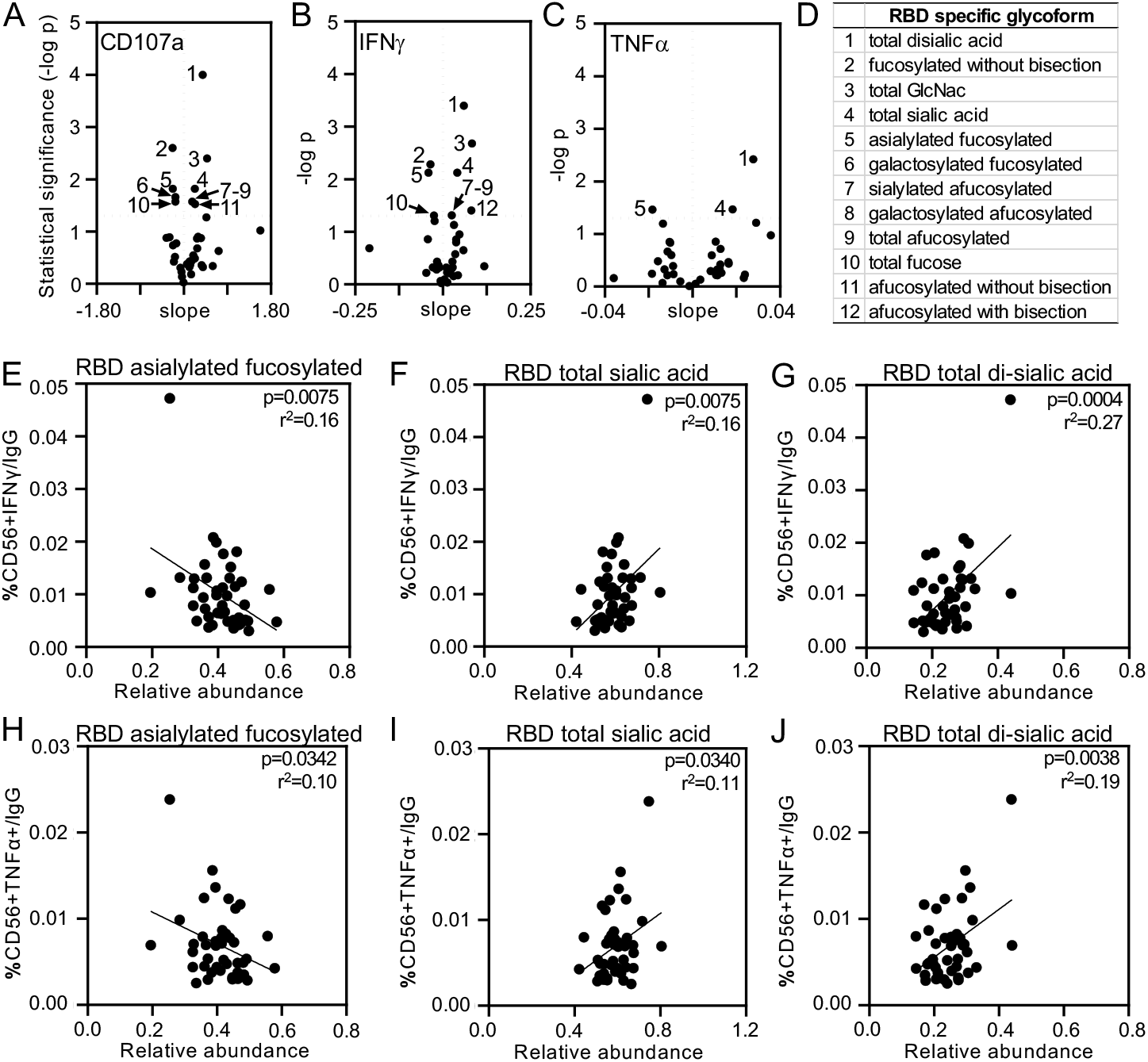
Differential fucose and sialic acid on vaccine specific IgG link FcγRIIIa/CD16a mediated IFNγ and TNFα production. Volcano plots depict slope and statistical significance (-log p) from linear regression assessing the dependency of receptor binding domain (RBD) ADNKA by (A) % CD56 CD107a, (B) IFNγ and (C) TNFα on different RBD specific IgG glycans. (D) Relationships where p<0.05 are enumerated and identified. Data for antibody dependent natural killer cell activation (ADNKA) markers of IFNγ (middle row) and TNFα (bottom row) per RBD IgG and relative abundance of RBD specific (E and H) asialylated fucosylated, (F and I) total sialic and (G and J) total di-sialic acid are plotted. Statistical significances were evaluated by linear regression.

**Supplemental Figure 5:**
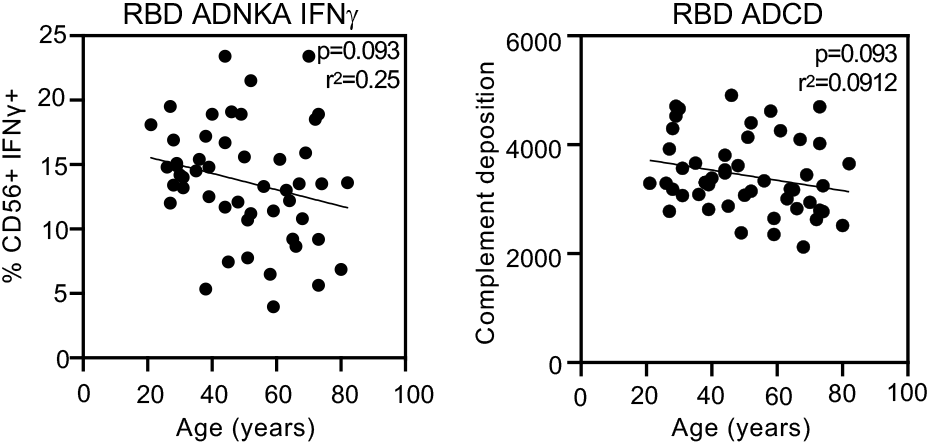
Minimal relationship between age and vaccine specific antibody dependent complement deposition (ADCD). Receptor binding domain (RBD) specific C3 deposition and age are plotted (right panel). The relationship between age and RBD antibody dependent natural killer cell activation (ADNKA) as measured by IFNγ are shown (left panel). Linear regression with p value adjusted for sex is reported.

**Supplemental Figure 6:**
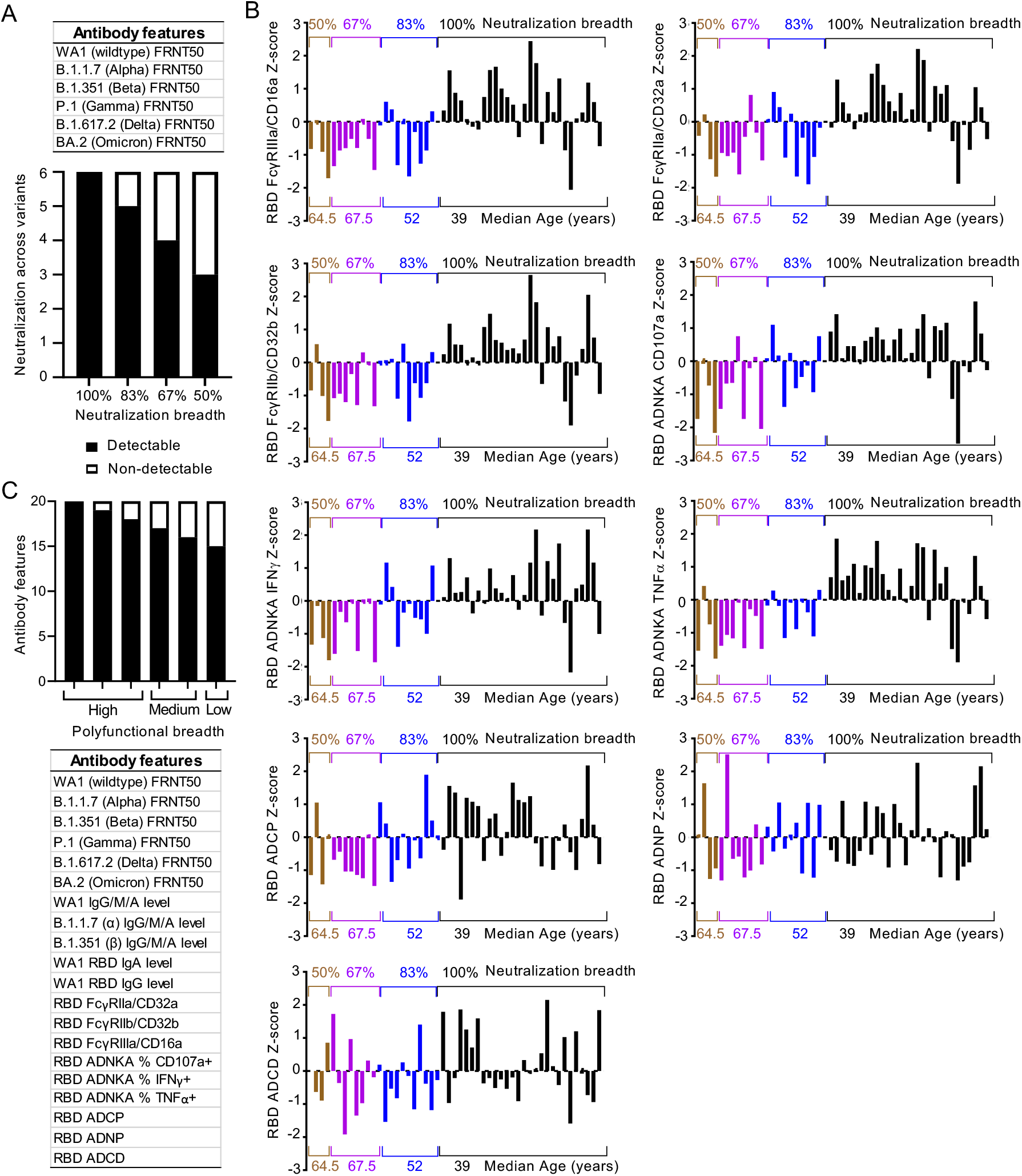
Antibodies function by the combination of Fab and Fc domains. (A) Neutralization breadth across all 6 SARS-CoV-2 wildtype and clinical variants was calculated for each individual. In this cohort, individual responses fell into four main categories: those with detectable neutralizing activity for 100% of viruses tested, 83%, 67% and 50%. (B) Histograms depict the Z scored data for each vaccine specific Fc effector function tested. Each column represents one individual. Groupings are by neutralization breadth categories described in (A). (C) Vaccine specific polyfunctional breadth was calculated for each individual with all 20 vaccine specific features listed. In this cohort, individual responses fell into three main categories: those with high (90-100%), medium (80-90%) and low (<80%) of functions detected.

